# The Community Opinions on Vaccine Issues and Decisions (COVID) Survey: Using a rapid Knowledge, Attitude and Practice (KAP) survey in supporting a community engagement approach to address COVID-19 vaccine uptake initiatives

**DOI:** 10.1101/2021.04.11.21255260

**Authors:** Najeeb Rahman

**Affiliations:** Leeds Teaching Hospitals NHS Trust

**Keywords:** Attitudes, Community, COVID-19, Ethnicity, Islam, Knowledge, Minority, Mosque, Practice Vaccine

## Abstract

**Background:** In the UK, it is acknowledged that ethnic minority communities have lower vaccination uptake rates compared with their White ethnic counterparts. It is additionally recognised that the UK Muslim community represents diverse ethnicities, which is reflected through their places of worship, namely mosques. Given the current pandemic and the rollout of the COVID-19 Vaccination campaign, mosques, in their capacity as community organisations, have been involved in health promotion initiatives such as webinars. The objective of this project was to support and improve COVID-19 Vaccine related health promotion activities being delivered by mosques in the UK city of Leeds by using a rapidly administered KAP survey.

**Methods:** A short survey was developed, reviewed for appropriateness by relevant mosque leaders, and distributed electronically to 2 mosque congregations. Analysis involved cumulative average scores for key domains with adjustment for key demographics, as well as a review of engagement preferences. Findings were fed back during an engagement webinar hosted by one of the mosques.

**Results:** There were a total of 151 participants, majority were male (95), between the ages of 31-50 (88) and of Asian ethnicity (102). Average Knowledge, Attitude and Practice scores for participants from Leeds Grand Mosque were 67,69 and 74% respectively, with participants from Leeds Makkah Mosque scoring 65, 62 and 67% respectively. Female participants as well as those aged less than 30 years scored poorer across all domains compared with the group average. The most important sources of information in relation to the vaccine were considered to be General Practitioners (GP) and National Health Service (NHS) or Government Websites.

**Conclusions:** A KAP survey is a useful tool to develop insights on community perspectives to the COVID-19 Vaccine, and can be rapidly deployed through community organisations such as mosques. Survey findings can then be used to improve the nature of health promotion, community engagement and service delivery in relation to mosques and their congregations. Adapting the survey for other groups and communities, as well as scaling up the partnership-based approaches to survey administration would support the development of multi-component strategies to address vaccine concerns and uptake.

## Background

Minoritised ethnic communities are less likely to be vaccinated than majority White groups [1]. A range of barriers have been considered including perception of risk, low confidence in the vaccine, distrust, access barriers, inconvenience, socio-demographic context and lack of endorsement, lack of vaccine offer or lack of communication from trusted providers and community leaders. Community engagement is key to responding to issues of vaccine hesitancy [2]. Within the Muslim community, a number of initiatives, in particular webinars and social media flyers, have been used to try and address some of the concerns [3]. Muslims in the UK represent diverse ethnic groups, and as such, organisations such as mosques provide a lens into such communities, rendering them suitable hubs to explore health and social issues [4]. However, there remains limited opportunities for communities to share their opinions in a non-stigmatised way so that initiatives can be tailored more appropriately. While health promotion activities have been implemented through UK mosques, they represent an underutilised opportunity [5,6,7]. In recognising that mosques are often considered an area of community activity, of which a number were hosting COVID-19 related webinars, it follows that such sites could serve as a means of enhancing community engagement and participation by seeking the opinions of mosque congregations to help inform webinar planning and discussion, as well as planning considerations for any future initiatives. It also follows that such engagement could support the development of improved community insights on the diversity and challenges of specific issues within the framework of the vaccine hesitancy model (inclusive of the ‘3C’s of Confidence, Complacency and Convenience), and so better guide support and interventions [8]. One way of exploring community health behaviours is by utilising Knowledge, Attitude Practice (KAP) surveys [9]. In addition, an emphasis on rapidity is required given the dynamic circumstances presented by the pandemic, which for all intents and purposes, represents a humanitarian crisis given its large geographical area of involvement, significant economic resource burden, and impact on ensuring fundamental human rights [10]. The aim of this project was to conduct a rapid KAP survey on behalf of participating Mosques to provide insights to support improvements in the planning and delivery of vaccine related initiatives relevant to their congregations and hence local community.

## Methods

### Study Setting

Two mosques, Leeds Grand Mosque and Leeds Makkah Mosque were purposively selected in view of their ongoing participation in community events such as webinars, engaged mosque leadership and located in the same postcode. The author additionally is known to the leadership of both mosques, and a member of the congregation of Leeds Grand Mosque, and so the prerequisite degree of trust and mutual understanding for any community partnership project was in place.

### Sample size

The usual ‘Juma’ah’ Friday prayer congregation size of both these mosques was between 1000-1200 worshipers prior to the pandemic, with regular daily prayer congregations between 30-60, and with both mosques having active Facebook pages and YouTube channels. Given that the survey would be distributed electronically, and recognising the engagement with social media posts can be variable, as well as that this was a feasibility study exploring the approach adopted rather than demonstrating true cohort representation, a formal sample size was not calculated.

### Questionnaire

An online survey (using Googleforms) was used to develop a short Knowledge Attitude Practice survey comprising of 5 sections and a total of 25 questions. The survey questions are outlined in an additional file [see Additional File 1], however in summary:

- Section A consisted of 6 questions focused on knowledge-based aspects
- Section B had 4 questions exploring attitudes
- Section C contained 5 questions examining behaviours
- Section D had 3 questions soliciting ideas and preferences on future activities and participation barriers
- Section E had 7 questions focusing on demographics, sources of information and an opportunity to offer further comments

Knowledge, attitude and practice questions were structured using a 5-point Likert-type scale of Strongly Disagree to Strongly Agree, as well as a separate ‘Don’t know option. Questions included both positive and negative framing, where in positive questions, Strongly Disagree correlated with a score of 1, and Strongly Agree with 5, with the inverse being the case with negatively framed questions. Don’t know (or absent) responses scored 0. Ethical considerations on the survey design are outlined in the ethics section below.

### Validation

The mosque committee or Imam were consulted to review suitability and appropriateness of questions prior to electronic distribution. A key element was to ensure the balance of brevity to support the mosques’ role in disseminating and encouraging participation with the survey, as well as relevance of topics, which were based on themes which the mosque leadership were aware of as issues within the community.

### Survey distribution

A social-media flyer suitable for WhatsApp, Facebook and email distribution was developed for each mosque, with a unique GoogleForms link and QR code for each mosque. The mosque committee or Imam were asked to facilitate electronic distribution of the survey through the relevant mosque social media channels. Informal agreements were made with the relevant mosque committee or Imam to support electronic distribution of the survey with relevant reminders to support completion, as well as to commit to a feedback meeting to discuss the results and findings. The surveys were open for collecting responses for between 1-2 weeks dependant on the preferences of each mosque during the second half of January 2021.

### Data and Analysis

Data was stored on a shared Google Drive with limited access to the author and mosque committee leaders or Imam only. Results were exported onto Googlesheets, where, following removal of duplicate entries, a numerical value was allocated to each of the responses, with average cumulative scores then calculated for each question, as well as for each domain of knowledge, attitude and practice. Domain averages were additionally explored by demographic characteristics to provide a descriptive analysis of the survey sample. For each domain, average scores of less than or equal to 50% were categorized as poor in terms of overall health behaviour, with 51-69% being adequate, and greater than 70% representing good. Free texts statements and comments were reviewed for general themes and ideas to feedback to the mosque leadership, although formal thematic analysis was not performed.

## Results

A total of 90 participants responded to the survey run at Leeds Grand Mosque (LGM), with 61 participants from Leeds Makkah Mosque (LMM). The majority of participants were male, aged between 31 and 50 years, and of Asian origin as summarised in Table 1 below. While data for further age and ethnicity subgroups were collected (as outlined in the sample survey), these were compiled into larger subgroups for ease of analysis.

**Table 1:** Basic demographic characteristics of participants

| Demographic characteristics | LGM (%) | LMM (%) |
| --- | --- | --- |
| Total Respondents (n) | 90 | 61 |
| Sex<br>Male<br>Female<br>Not declared | 61 (67.8)<br>29 (32.2)<br>- | 34 (55.7)<br>19 (31.1)<br>8 (13.1) |
| Age<br>16-30 years<br>31-50 years<br>51-70 years<br>Not declared | 22 (24.4)<br>50 (55.6)<br>18 (20)<br>- | 10 (16.4)<br>38 (62.3)<br>11 (18)<br>2 (3) |
| Ethnicity<br>Asian<br>Arab<br>White<br>Black<br>Other<br>Not declared | 45 (50)<br>23 (25.6)<br>11 (12.2)<br>4 (4.4)<br>5 (5.6)<br>2 (2.2) | 57 (93.4)<br>4 (6.6) |

With regards to the knowledge domain questions, cumulative averages demonstrated adequate scores at 67% and 65% for LGM and LMM respectively, Males and those aged greater than 51 years generally displayed ‘good’ or more positive health behaviours than those who were female or below the age of 31. In relation to LGM, participants of Arab and White ethnicity were categorised as having good knowledge compared with their Asian and Black counterpart. These findings are summarised in Table 2.

**Table 2:** Summary of Knowledge Domain scores for the total sample and key groups.

| Knowledge Domain | LGM | LMM |
| --- | --- | --- |
| Cumulative Score (%) | 67 | 65 |
| Male | 70 | 68 |
| Female | 62 | 63 |
| Age<br>16-30 years<br>31-50 years | 66<br>67 | 61<br>65 |
| • 51-70 years | 71 | 69 |
| Ethnicity |  |  |
| • Asian | 65 | 68 |
| • Arab | 73 | - |
| • White | 72 | - |
| • Black | 56 | - |

When considering attitudinal aspects towards the COVID-19 vaccine, again cohorts of both mosques scored adequately with participants from LGM more positively skewed at 69% compared with 62% for participants from LMM. Table 3 outlines the scores in relation to the attitude domain. Similar trends in reference to sex and age were noted as in knowledge domain scores, but did not follow through when considering ethnicity, with Black respondents demonstrating ‘good’ attitude with scores of 84%.

**Table 3:** Summary of Attitude Domain scores for the total sample and key groups.

| Attitude Domain | LGM | LMM |
| --- | --- | --- |
| Cumulative Score (%) | 69 | 62 |
| Male | 70 | 67 |
| Female | 67 | 61 |
| Age |  |  |
| • 16-30 years | 65 | 54 |
| • 31-50 years | 67 | 63 |
| • 51-70 years | 78 | 66 |
| Ethnicity |  |  |
| • Asian | 69 | 65 |
| • Arab | 68 | - |
| • White | 69 | - |
| • Black | 84 | - |

Cohorts of both mosques scored most positively in terms of practice preferences, with LGM participants categorised as good, with 74%, and LMM as adequate with 67%. In relation to LGM, these positive trends were again more marked in males, those older than 51, as well as in participants of Black ethnicity. When considering LMM, males again scored more highly when compared with females, however when considering age, those aged greater than 51 scored lower than those aged 31-50, which is a different correlation when considering knowledge and attitude across the same cohort. The summary data can be reviewed in Table 4.

**Table 4:** Summary of Practice Domain scores for the total sample and key groups.

| Practice Domain | LGM | LMM |
| --- | --- | --- |
| Cumulative Score (%) | 74 | 67 |
| Male | 76 | 73 |
| Female | 70 | 66 |
| Age |  |  |
| • 16-30 years | 70 | 61 |
| • 31-50 years | 73 | 70 |
| • 51-70 years | 81 | 66 |
| Ethnicity |  |  |
| • Asian | 75 | 70 |
| • Arab | 71 | - |
| • White | 74 | - |
| • Black | 85 | - |

The survey offered the opportunity for participants to provide suggestions on how to improve engagement or additional comments on their experiences of the vaccine roll-out. Table 5 summarises a sample of these which represent common themes around improving confidence in, and improving accessibility and convenience of obtaining the vaccine, as well as addressing issues of complacency through faith-based and contextualised approaches.

**Table 5:**
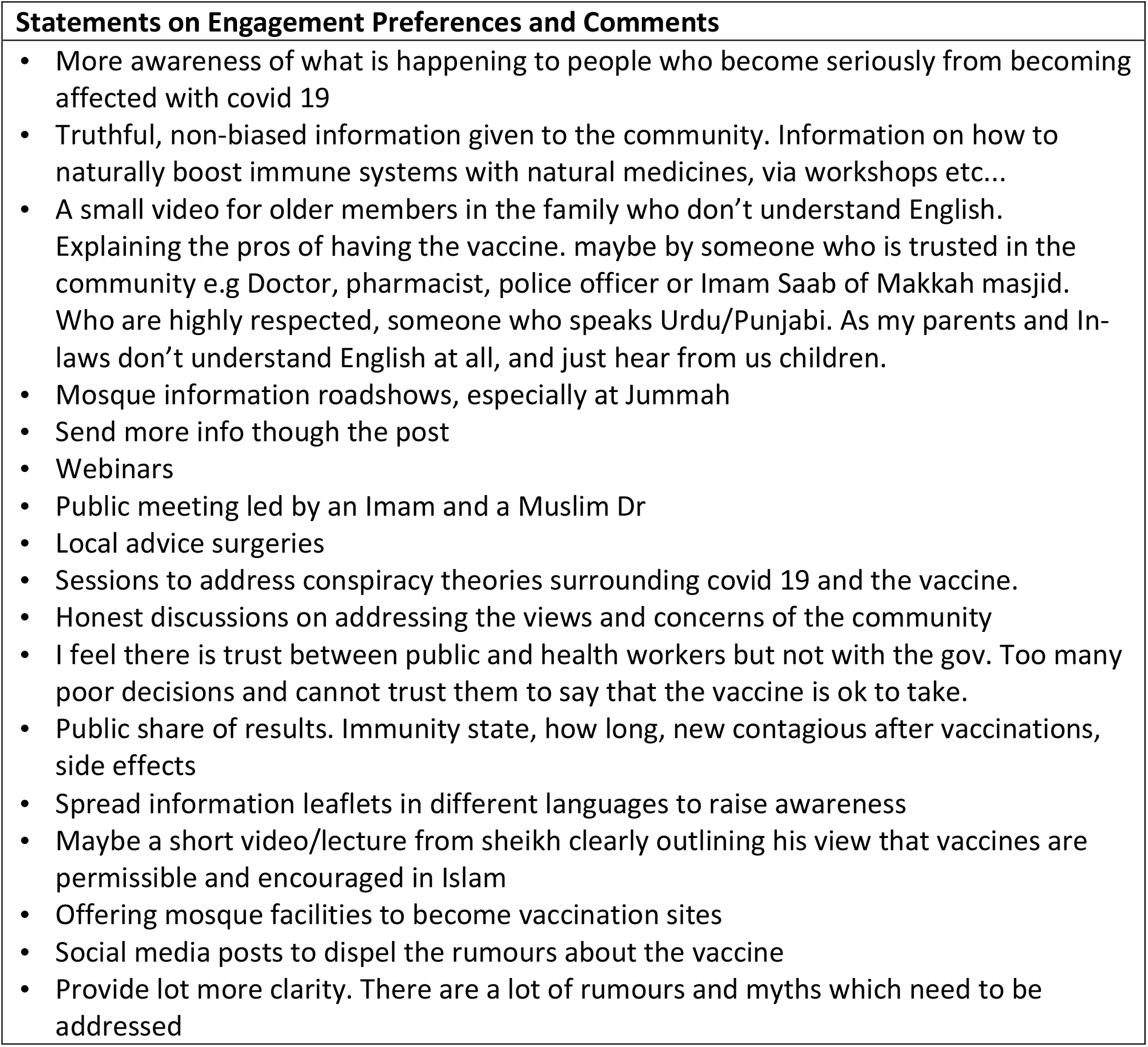
Sample of statements provided on engagement preferences and comments.

When considering sources of information, the most important were considered to be General Practitioners (GP) and NHS or Government Websites, which accounted for the most number of responses for participants of both mosques. Table 6 summarises the range of information sources for the group when as to select the three most important ones used to support COVID19 and vaccine related decisions.

**Table 6:**
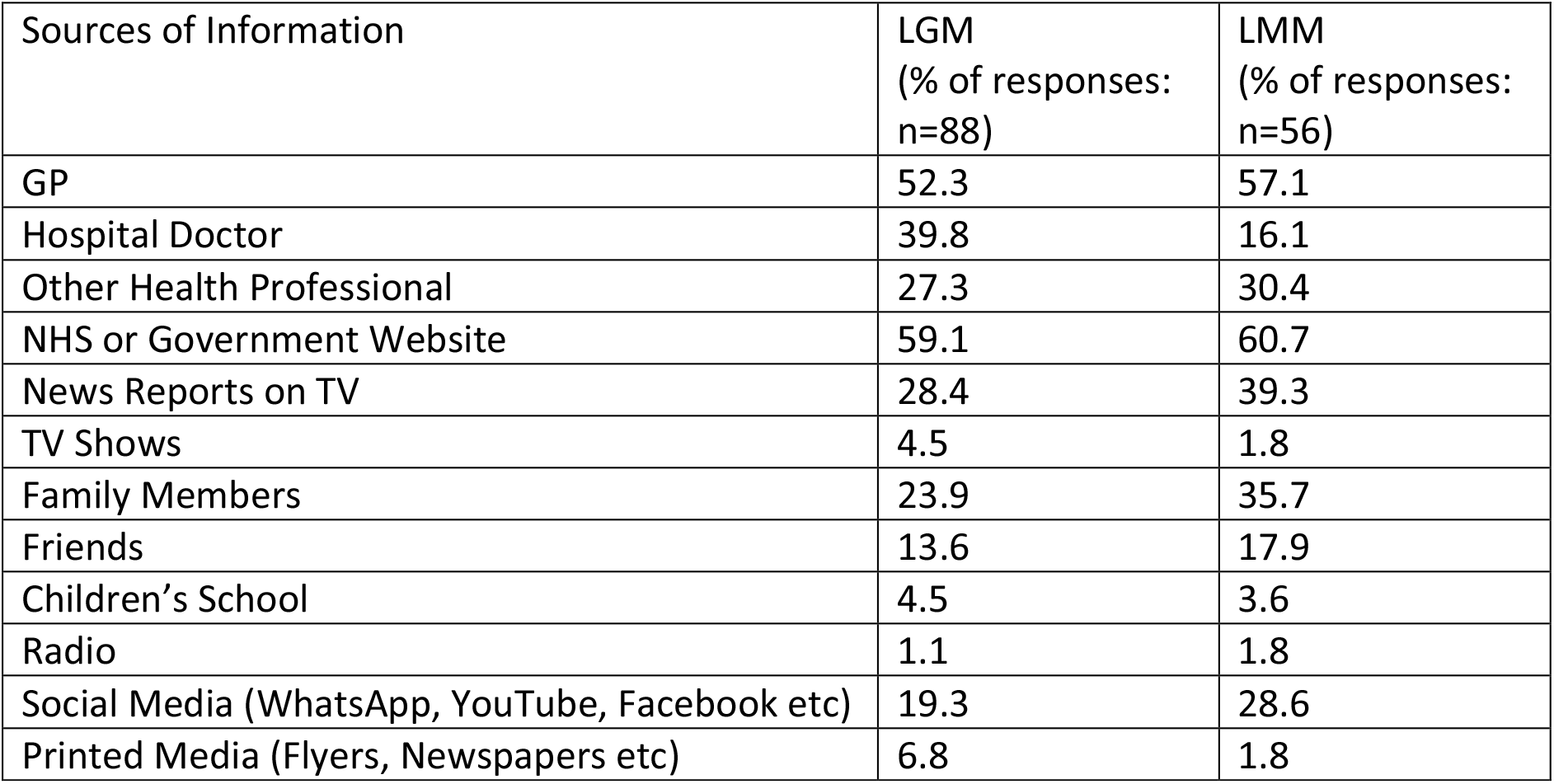
Summary of most important sources of information in making decisions regard COVID-19 and the vaccine.

| Sources of Information | LGM<br>(% of responses:<br>n=88) | LMM<br>(% of responses:<br>n=56) |
| --- | --- | --- |
| GP | 52.3 | 57.1 |
| Hospital Doctor | 39.8 | 16.1 |
| Other Health Professional | 27.3 | 30.4 |
| NHS or Government Website | 59.1 | 60.7 |
| News Reports on TV | 28.4 | 39.3 |
| TV Shows | 4.5 | 1.8 |
| Family Members | 23.9 | 35.7 |
| Friends | 13.6 | 17.9 |
| Children's School | 4.5 | 3.6 |
| Radio | 1.1 | 1.8 |
| Social Media (WhatsApp, YouTube, Facebook etc) | 19.3 | 28.6 |
| Printed Media (Flyers, Newspapers etc) | 6.8 | 1.8 |

## Discussion

The results show that in general, positive practice behaviours are observed, despite knowledge gaps and guarded attitudes towards vaccination. This is an important and reassuring observation and contrasts with the current more worrying perspectives on vaccine uptake in minority communities stated to be in the region of 57% willingness to take the vaccine [11]. Furthermore, the findings suggest that a range of other engagement approaches may be needed to address variations observed for younger people as well as females. An interesting point is when considering perspectives of Black participants, where despite adequate (trending towards poor) knowledge, the cohort, although a small sample, demonstrated good attitude and practice. Such a stark variation was not observed amongst other ethnicities, and as such, warrants further exploration. When comparing the two mosques, despite being only 0.6miles away from each other, clearly represent different communities in terms attitudes and practice, with the variation more marked amongst women and people below the age of 31. A review of important sources of information demonstrated the trust placed in official NHS and government sources as well as GPs.

However, there were differences in the two congregations when considering other sources, with News Reports, Family, Friends and Social media featuring more highly for participants of Leeds Makkah Mosque. Participants from Leeds Grand Mosque appeared to have greater access to other health professionals and hospital doctors, as these featured more highly when compared to their Makkah Mosque counterparts. Interestingly, and requiring further exploration, information via schools, radio and printed media was rated uniformly low for both groups, which may either represent missed opportunities to enhance communication and messaging, or perhaps reflects the general lack of utility of such sources. These findings further demonstrate the need for localized and tailored interventions to address health behaviour variations [12]. Within the 3Cs model, comments illustrated the need to regain trust in the health system, as well as to address access barriers with facilitation support. A key aspect was the value placed in community leaders and health professionals in supporting positive health behaviours, which reinforces the view that narrowing the gap between communities and health workers in terms of accessibility would serve to address some of the barriers related to vaccine uptake. Such measures may include support to develop and strategically embed a range of culturally competent and inclusive multi-component engagement tools which can be modelled on existing health promotion strategies, such as social media and school engagement, Very Brief Advice and Group Consultations [13,14,15]. In addition, targeted health promotion aligned with culturally competent concepts may be of additional value, for example, utilizing the Islamic principles of the preservation of life and protection of society in challenging the complacency of individuals [16]. Adapting the survey for other communities and faith groups, as well as administering it in other Muslim community settings would yield further insights. A key consideration would be the additional capacity required to support researchers and project teams (ideally involving members of the places of worship so as to support partnership working and trust) to validate questions, review results, and support the development and implementation of interventions.

## Limitations

Key limitations of this study include the fact it was conducted in English, and this may not have been the preferred language of participants. In addition, the wording and style of the survey had not been piloted with a representative sample beyond the mosque leadership prior to distribution. The option of electronic distribution would exclude those congregation members who are not supported with digital access. In addition, a significant proportion of mosque attendees are male, and hence it is important to acknowledge the inequitable community representation of females when conducting such studies through mosques. Much of these limitations could be addressed through a more engaged partnership-based approach with Mosques, other places of worship, and community-based organisations, supported with increased capacity and resources to conduct such activities, including further categorisation of demographics to include deprivation metrics.

## Conclusions

The results of this project demonstrate the feasibility in engaging with community-based organisations such as mosques in exploring community health behaviours. The findings demonstrate the importance of localised approaches in addressing the 3Cs model on vaccine hesitancy, as well as confirming that involving local leaders and health care professionals through webinars are consistent with engagement preferences. The COVID Survey represents a rapid, pragmatic, adaptable and scalable model in developing community partnerships to support local vaccine promotion, as well as local insights to support future policy planning and implementation decisions to ensure a more equitable approach to vaccine uptake. Importantly, it reinforces the value in localised and community-focused approaches in responding to health and wellbeing as well as social and welfare needs, which will be of increasing significance in relation to the ongoing and unequal demands of the COVID-19 pandemic.

## Supporting information

Additional File 1_COVID KAP Survey

## Data Availability

The datasets used and/or analysed during the current study are available from the corresponding author on reasonable request.

## List of Abbreviations

KAP: Knowledge Attitude Practice
COVID-19: Coronavirus Disease 2019
GP: General Practitioner
NHS: National Health Service
3Cs: Confidence, Complacency, Convenience
QR code: Quick Response code
LGM: Leeds Grand Mosque
LMM: Leeds Makkah Mosque
HRA: Health Research Authority
mRNA: Messenger ribonucleic acid

## Declarations

### Ethics Approval and consent to participate

This project was considered a public survey to help improve the health promotion service offering of the participating mosques in their role of providing COVID-19 related information. Having followed the Health Research Authority (HRA) decision tool and research definition guidance (located via their website at http://www.hra-decisiontools.org.uk/research/index.html), it was concluded that this survey did not constitute research, and as such, formal ethics review was not sought. In addition, advice was sought from the research governance office of the Research and Innovation Department at Leeds Teaching Hospitals NHS Trust, who also agreed that ethical review would not be required. However, ethical conduct principles were adhered to as follows:

- Study information was provided, as well as electronic informed consent was obtained, prior to survey completion.
- No personal identifiers were collected, and participation was voluntary, with the option to withdraw responses by contacting the study author or mosque committee by email.
- No personal health data was collected
- There were no compulsory questions, and participants had the option not to declare characteristics of sex, age and ethnicity.
- Data was stored on a secure, password protected drive with access only to the author and mosque leadership.
- Terms of references were outlined to ensure that the author and mosque leadership understood the nature of the partnership in conducting the survey, the analysis and supporting the feedback of results.

## Consent for publication

Informed consent to publish was obtained at time of survey implementation

## Author’s Contribution

NR was the sole author of this manuscript.

## Acknowledgements

The author would like to thank the Leeds Grand Mosque Committee Chair, Dr Ihab Ibrahim as well as Mr Lasaad Laouini who supported the distribution of the survey and included the feedback as part of an engagement webinar. In addition, the author would like to acknowledge Imam Qari Muhammed Asim of Leeds Makkah Mosque for his guidance and support in ensuring survey distribution to the mosque congregation.

## Author’s information

Najeeb Rahman is a Consultant in Emergency Medicine, but has additional skills and expertise in humanitarian response, global health and development through roles with the medical charity, Doctors Worldwide. In addition, he is a member of the Royal College of Emergency Medicine’s Global Emergency Medicine Committee, and Emergency Medicine and Public Health Special Interest Group.

