## Additional File 1_COVID KAP Survey for "The Community Opinions on Vaccine Issues and Decisions (COVID) Survey: Using a rapid Knowledge, Attitude and Practice (KAP) survey in supporting a community engagement approach to address COVID-19 vaccine uptake initiatives"

### COVID Survey

Community Opinions on Vaccine Issues and Decisions (COVID):
A short KAP survey of the Muslim Community attending Leeds Grand Mosque

#### What is this survey about?

-We know that ethnic minority groups have been affected badly the COVID19 pandemic.
-We also know that within the Muslim community, a number of initiatives, in particular webinars and social media flyers, have been used to try and address some of the concerns, particularly around vaccines.
-However, there have been limited chances for the community to share their opinions to help guide that kind of events they would like to see and be involved in.
-This is your chance to share your views on issues of the COVID19 vaccine.
-This short survey is voluntary, and will take about 5 mins to complete.
- Please do try and complete all the questions. Thank you

#### What is involved?

The survey will take about 5 mins to complete.
It consists of 5 short sections which includes check boxes, as well as the chance for you to share some comments and tell us a bit about yourself.

#### Who is carrying out this survey?

- The survey is being conducted by Dr Najeeb Rahman, who is an Emergency Medicine consultant based at Leeds Teaching Hospitals NHS Trust, is a trustee of the medical charity Doctors Worldwide, and is also the founder of the Frontline Collaboration Against COVID-19. This initiative brings together doctors and humanitarians to better respond the COVID19 pandemic.
- This survey is being conducted on behalf of the group and in partnership with community based organisations so as to better understand the needs and opinions of the Muslim community in particular.
- This version of the survey is being conducted in partnership with Leeds Grand Mosque.
- Findings may be shared in the public domain through reports and publications so as to raise awareness of issues and concerns that the community is facing.

#### Will my details be kept confidential?

YES. The survey is anonymised, and no personal identifiers are being collected. All data will be kept secure and confidential in line with the Data Protection Act.

#### Can I withdraw from this survey? What if I have a query

#### Thank you for taking part in this survey, and helping to improve our understanding of the community. This would not be possible without your support and involvement.

#### Consent

- I confirm that I have reviewed the information regarding this survey
- I understand that my participation is voluntary and that I am free to withdraw at any time without giving any reason.
- I agree for my responses in this questionnaire to be used for the purpose of this survey study, including any publications or projects, only and according to the Data Protection Act, meaning that any personal information is kept strictly confidential.

**- By continuing on with this survey, I effectively consent and agree to participate in this assessment.**

#### Section A: Your understanding of vaccines

The following questions refer to your understanding of the Pfizer-BioNTech, Oxford AstraZeneca and Moderna Vaccines.
Please tick the box which most accurately reflects your views on the following statements.

1. 1. The current approved COVID-19 vaccines are safe for use in the general population.

*Mark only one oval.*

- Strongly Agree
- Agree
- Neither Agree or Disagree
- Disagree
- Strongly Disagree
- Don't Know

2. 2. From an Islamic perspective, vaccines are considered permissible and halal by the majority of Islamic scholars.

*Mark only one oval.*

- Strongly Agree
- Agree
- Neither Agree or Disagree
- Disagree
- Strongly Disagree
- Don't Know

3. 3. The severity of COVID-19 illness if you get sick is the same regardless if you have had the vaccine or not.

*Mark only one oval.*

- Strongly Agree
- Agree
- Neither Agree or Disagree
- Disagree
- Strongly Disagree
- Don't Know

4. 4. Vaccines are necessary, and one of the only ways to help return back to a more normal way of life.

*Mark only one oval.*

- Strongly Agree
- Agree
- Neither Agree or Disagree
- Disagree
- Strongly Disagree
- Don't Know

5. 5. The vaccines are clearly linked to many serious, life-changing side effects

*Mark only one oval.*

- Strongly Agree
- Agree
- Neither Agree or Disagree
- Disagree
- Strongly Disagree
- Don't Know

#### Section B: Your thoughts on the vaccine

The following questions refer to your thoughts on the Pfizer-BioNTech, Oxford AstraZeneca and Moderna Vaccines.
Please tick the box which most accurately reflects your views on the following statements.

6. 6. Using mRNA technology is a safe and effective method of developing new vaccines.

*Mark only one oval.*

- Strongly Agree
- Agree
- Neither Agree or Disagree
- Disagree
- Strongly Disagree
- Don't Know

7. 7. It is better to delay myself or my family getting the vaccine to wait and see what happens with the general public as the vaccine is rolled out.

*Mark only one oval.*

- Strongly Agree
- Agree
- Neither Agree or Disagree
- Disagree
- Strongly Disagree
- Don't Know

8. 8. The benefits of vaccines outweigh any risks or side effects.

*Mark only one oval.*

- Strongly Agree
- Agree
- Neither Agree or Disagree
- Disagree
- Strongly Disagree
- Don't Know

9. 9. It is better to gain immunity through getting sick with COVID19 instead of using the vaccine.

*Mark only one oval.*

- Strongly Agree
- Agree
- Neither Agree or Disagree
- Disagree
- Strongly Disagree
- Don't Know

10. 10. I trust the information and guidance being provided by the Government, NHS and health professionals

*Mark only one oval.*

- Strongly Agree
- Agree
- Neither Agree or Disagree
- Disagree
- Strongly Disagree
- Don't Know

#### Section C: Your preferences and practice

The following questions refer to your preferences regarding the Pfizer-BioNTech, Oxford AstraZeneca and Moderna Vaccines.
Please tick the box which most accurately reflects your views on the following statements.

11. 11. If myself or someone in my family are offered to take the vaccine, I will refuse.

*Mark only one oval.*

- Strongly Agree
- Agree
- Neither Agree or Disagree
- Disagree
- Strongly Disagree
- Don't Know

12. 12. I am able to easily discuss my concerns about the vaccine with my GP or other health professional (such as a pharmacist).

*Mark only one oval.*

- Strongly Agree
- Agree
- Neither Agree or Disagree
- Disagree
- Strongly Disagree
- Don't Know

13. 13. I usually take other vaccines when appropriate, and encourage my family members do to the same (such as with Flu vaccine, or childhood school vaccinations)

*Mark only one oval.*

- Strongly Agree
- Agree
- Neither Agree or Disagree
- Disagree
- Strongly Disagree
- Don't Know

14. 14. I trust and follow the advice and rulings that Islamic Scholars and Imams have issued in relation to the vaccine.

*Mark only one oval.*

- Strongly Agree
- Agree
- Neither Agree or Disagree
- Disagree
- Strongly Disagree
- Don't Know

15. 15. I will follow the advice and recommendations from my GP/Hospital Doctor if I or my family members are invited to receive the vaccine.

*Mark only one oval.*

- Strongly Agree
- Agree
- Neither Agree or Disagree
- Disagree
- Strongly Disagree
- Don't Know

#### Section D: Planning Ahead

16. 16. Please tell us what kind of activities you would like to see arranged to help provide information or address any concerns about COVID19 and related issues.

17. 17. What do you feel stops you being involved in such activities?

18. 18. What do you think we will improve participation and involvement of the community in such activities?

#### Section E: About Yourself

19. 19. What is your gender?

*Mark only one oval.*

- Female
- Male
- Prefer not to say

20. 20. What is your age?

*Mark only one oval.*

- < 16 years
- 16-20 years
- 21-30 years
- 31-40 years
- 41-50 years
- 51-60 years
- 61-70 years
- >70 years

21. 21. What is your ethnicity?

*Mark only one oval.*

- White/White British
- White/Eastern European
- White/Other
- Middle Eastern/Arab
- Middle Eastern/Non-Arab
- Asian or Asian British/Indian
- Asian or Asian British/Bangladeshi
- Asian or Asian British/Pakistani
- Asian or Asian British/Other
- Black or Black British/Caribbean
- Black or Black British/West African
- Black or Black British/East African
- Black or Black British/Other
- Other Ethnic Group

22. 22. Please select the 3 most important sources of information that help you to make decisions regarding COVID19 and the vaccine.

*Check all that apply.*

- [ ] GP
- [ ] Hospital Doctor
- [ ] Other Health Professional
- [ ] NHS or Government Website
- [ ] News Reports on TV
- [ ] TV Shows
- [ ] Family Members
- [ ] Friends
- [ ] Children's School
- [ ] Radio
- [ ] Social Media (WhatsApp, YouTube, Facebook, Twitter etc)
- [ ] Printed Media (Flyers, Newspapers)

23. 23. Of your 3 selected options for the previous question, please give more details such as the name of the TV show, or name of the social media app or which family member.

24. 24. Which Mosque/Islamic Centre do you normally attend?

25. 25. Any other comments

Thank you for completing this survey

We are really grateful, as this survey could not have been done without your help.
If you would like to follow the progress of this survey including any outcomes, please or contact your local mosque/Islamic centre representative.

This content is neither created nor endorsed by Google.

Google Forms
